# Navigating the Privacy-Accuracy Tradeoff: Federated Survival Analysis with Binning and Differential Privacy

**DOI:** 10.1101/2024.10.09.24315159

**Authors:** Varsha Gouthamchand, Johan van Soest, Giovanni Arcuri, Andre Dekker, Andrea Damiani, Leonard Wee

## Abstract

Federated learning (FL) offers a decentralized approach to model training, allowing for data-driven insights while safeguarding patient privacy across institutions. In the Personal Health Train (PHT) paradigm, it is local model gradients from each institution, aggregated over a sample size of its own patients that are transmitted to a central server to be globally merged, rather than transmitting the patient data itself. However, certain attacks on a PHT infrastructure may risk compromising sensitive data. This study delves into the privacy-accuracy tradeoff in federated Cox Proportional Hazards (CoxPH) models for survival analysis by assessing two Privacy-Enhancing Techniques (PETs) added on top of the PHT approach. In one, we implemented a Discretized Cox model by grouping event times into finite bins to hide individual time-to-event data points. In another, we explored Local Differential Privacy by introducing noise to local model gradients. Our results demonstrate that both strategies can effectively mitigate privacy risks without significantly compromising numerical accuracy, reflected in only small variations of hazard ratios and cumulative baseline hazard curves. Our findings highlight the potential for enhancing privacy-preserving survival analysis within a PHT implementation and suggest practical solutions for multi-institutional research while mitigating the risk of re-identification attacks.

## Introduction

Federated learning (FL) has emerged as a groundbreaking class of decentralized approaches to training machine learning models and solving complex problems, particularly in scenarios where data sensitivity precludes traditional data-sharing methods [1]. The imperative to relocate or centralize privacy-sensitive data from its original sources is bypassed. Instead, the learning process occurs synchronously over multiple participating institutions or devices, sharing only individual results in the form of local model gradients, which are then merged into a global model. This approach has garnered significant attention and research interest across various sectors, like healthcare and medicine, but also extends to other domains [2]. As the digital landscape continues to evolve and data privacy regulations become increasingly stringent, FL stands out as a promising solution.

The Personal Health Train (PHT) [3-6], a manifesto implementing the FL principles, has recently seen significant traction [7-10]. This integrates several critical components to enable secure and privacy-preserving data analysis, including the technical, legal and governance aspects. Within this framework, “ Stations” serve as local data repositories where data is securely stored and accessed under predefined rules. “ Trains” refer to the algorithms or workflows that visit these stations to conduct data analysis locally at the data’s location, thereby bypassing the need to transfer individual-level data. The “ Track” or handler orchestrates the interactions between trains and stations, ensuring compliance with security protocols and facilitating the aggregation of results. The “ Aggregator” combines the local gradients from various stations, integrating distributed data sources while preserving privacy.

While this framework is designed to maintain data within its originating institution, recent studies have uncovered that FL methods are susceptible to specific attacks resulting in potential data leakage [11-16]. Kairouz et al. [17] have comprehensively analyzed the open challenges in FL and proposed potential solutions. Key threats identified include Unauthorized Access, where breaches in infrastructure security could expose sensitive information to unauthorized entities. Poisoning Attacks include data poisoning, where malicious actors introduce misleading data into the training process, and model poisoning, where harmful local updates are transmitted that compromise the integrity of the global model. Inference Attacks include membership inference attacks, where attackers can determine if specific data subjects were part of the training set by analyzing model updates, especially when sample sizes are limited, and model inversion attacks, where adversaries can reconstruct training data from model gradients, revealing sensitive information.

Recent research has explored implementing Cox Proportional Hazards (CoxPH) models within an FL framework [18], where data stations transmit subjects’ unique event times together with model gradients. However, this approach may potentially expose sensitive data to inference attacks, compromising data privacy. Brink et al. [19] demonstrated that the local model gradients shared by data stations for aggregation of the CoxPH model can lead to information leakage, particularly in cases where event times lack ties or censoring. This may make model inversion attacks easier since the federated Cox model sends repeated queries requesting local gradients until model convergence.

This paper investigates the tradeoff between privacy and accuracy in federated Cox analysis by experimenting with two Privacy-Enhancing Techniques (PETs). We build upon the implementation of federated Cox analysis as described by Lu et al. [18]. One of our proposed methods is discretized Cox analysis, where event times are grouped into bins by the participating data stations prior to fitting the local model, in the hope of increasing confidentiality in case of an adversarial attack. The other, we aim to address the vulnerability of sharing model gradients by incorporating Differential Privacy (DP) [20, 21]. This addresses inference risks by adding noise to model updates, ensuring that individual data points remain indistinguishable within the aggregated results [22]. Our main goal is to minimize privacy risks while maintaining the model’s overall accuracy. Lastly, we apply these methods to derive the cumulative baseline hazard curve in a federated setting and compare the results to the hazards estimated using the standard federated Cox results.

## Methods

### Data and infrastructure

For this study, three public datasets from The Cancer Imaging Archive (TCIA) - HN1 [23], HEAD-NECK [24], and OPC [25] - were used. Full details of the datasets can be found in the corresponding references. These datasets were reformatted to ensure a consistent nomenclature for compatibility with the federated algorithms used. The study used Vantage6 [26], an open-source federated framework based on the PHT approach. We leveraged the demo-network functionality of Vantage6, which allows users to set up a server along with the necessary data stations on the local machines, thereby simulating a test environment for enabling the execution and evaluation of user-defined tasks and algorithms. A visual representation of this setup is provided in *Fig 1*. The primary focus of this work is on the partial results (highlighted by red arrows) shared by the data stations for aggregation. The federated Cox algorithm used in this study had been previously adapted for the Vantage6 infrastructure [27]. It was further upgraded to be compatible with the latest version of Vantage6 (v4.5) for this work. The complete code for the algorithm can be accessed at https://github.com/MaastrichtU-CDS/v6-coxph/releases/tag/2.0.0.

**Fig 1.**
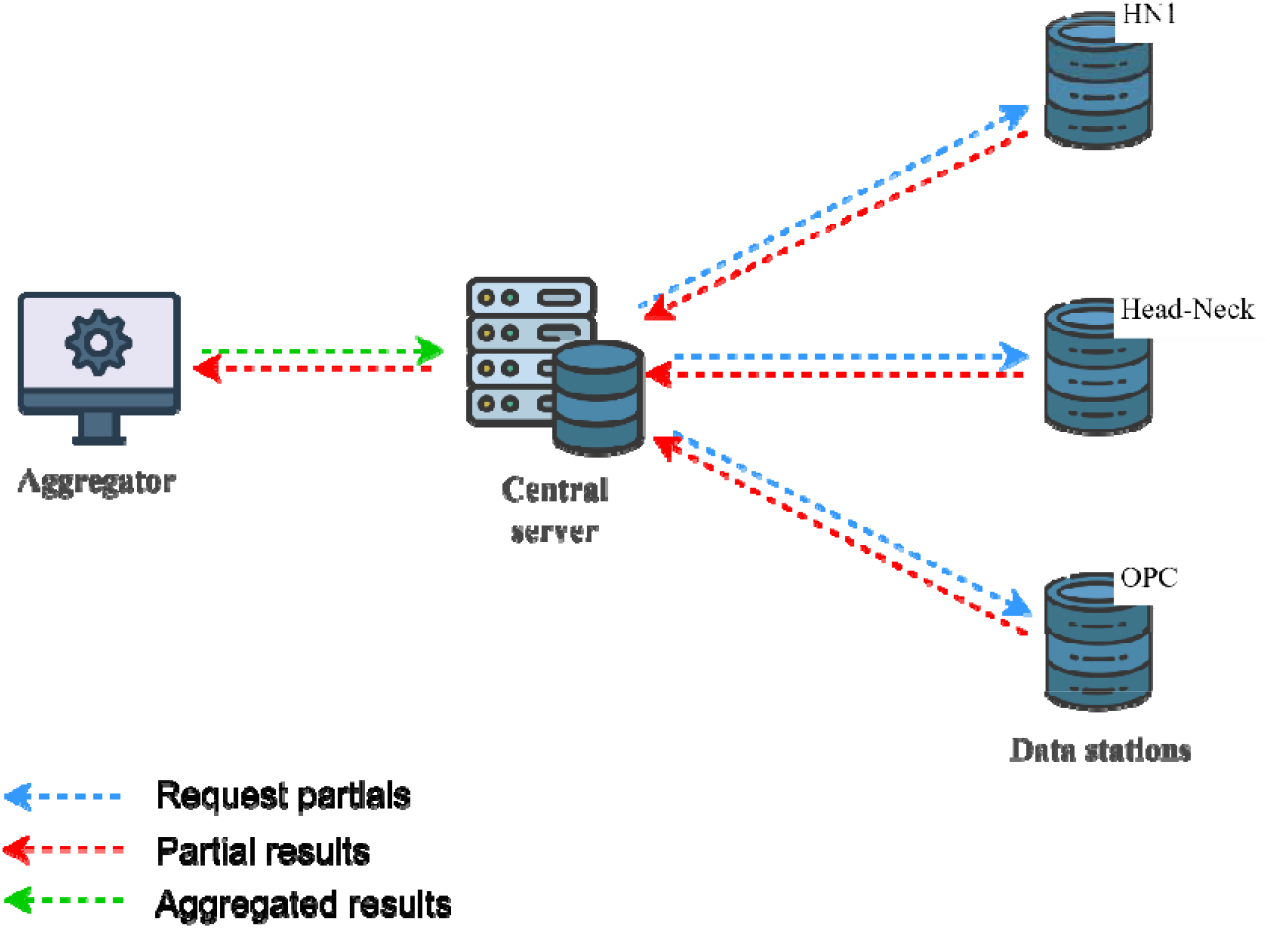
Visual illustration of Vantage6 setup used in this work

### Discretized Cox Analysis

Our initial experiment for federated Cox analysis involved implementing a discretized time approach rather than the standard continuous time method, which transmits unique event times from the institution as partial results. In the discretized method, event times are grouped into intervals before being sent to the central server for aggregation. Within this approach, we explored two binning strategies:

- **Fixed Binning**: Events are divided into equal-sized bins across all data stations based on optimal bin values determined beforehand.
- **Quantile Binning**: Events are categorized based on an optimal number of quantiles identified using the optimal bin values calculated beforehand by the algorithm. Quantile binning ensures that events are evenly distributed across each bin.

To determine the optimal number of bins for both strategies, we applied Sturges’ rule [28], defined as:

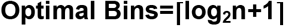

Where *n* represents the total sample size, and ∏ indicates rounding to the nearest integer.

For fixed bins, the stations were instructed to locally assign intervals using the “ histogram_bin_edges” function from the Python NumPy package, based on the optimal value. These intervals were shared with the server, which computed the global bin edges using the overall minimum and maximum values. For quantile bins, each station calculated the quantile edges for its data based on the optimal value, utilizing the “ quantile” function from the NumPy package. These quantile edges were shared with the server to determine the global edges.

The global edges in both scenarios were then sent back to the stations to compute the event counts for their respective bins, which were subsequently aggregated for the Cox analysis.

### Differential Privacy

Differential privacy (DP) reduces the risk of inference attacks by introducing noise either to the data or to the model updates, thereby increasing the likelihood that individual data points remain indistinguishable. Andreux et al., [29] discussed that introducing noise to local model gradients is sufficient to prevent attackers from reconstructing data. For this work, we applied Local Differential Privacy (LDP), where each data station perturbs its local gradients by introducing differential noise sampled from a Laplacian distribution before transmitting them to the server. This approach eliminates the need to trust the infrastructure’s integrity or rely solely on the server, as the noise is added directly at the data source. By doing so, the risk of inference attacks or the possibility of malicious actors posing as the server to extract private information is significantly reduced. In addition to perturbing the gradients, we also experimented with applying Laplacian noise to the input variables (model predictors) for 25% of the samples (patients) locally before the gradient computation at each data station.

The key privacy parameter in differential privacy, epsilon (ε), quantifies the extent of privacy loss by controlling the amount of randomness added to the data. A smaller epsilon value indicates stronger privacy, as it means less information about any individual data point can be inferred from the output. Epsilon provides a quantifiable way to balance the trade-off between data utility and privacy. To determine the optimal epsilon and the corresponding noise scale (sensitivity/epsilon), we conducted a series of Cox models using the three public datasets. We applied a combination of noise scales (the parameter ranging from 0.25 to 10) to the local gradients. The results were validated using a private dataset from Maastro - HN3, which includes 165 samples. The resulting Harrell’s concordance indices (C-Indices) are plotted in *Fig 3*.

### Ground Truth for Cumulative Baseline Hazards

Initially, for the standard federated Cox model, the Breslow estimator [30] was used to calculate the cumulative baseline hazard at each event time, which is defined as follows:

Where *d*_*i*_ represents the number of events at each unique time *t* and *X*β denotes the Linear Predictor (LP) scores.

### Cumulative Baseline Hazard for PETs

Following the derivation of the Cox model results for each PET experiment, we computed the cumulative baseline hazard using the Breslow estimator, either at each event bin edge (for binning) or at each event time (for DP), using the aggregated results from each dataset.

## Results

### Event Times Binning

The total sample size from the three public datasets consisted of 1,041 patients. The binning process began by requesting each data station to share its sample size. It was then aggregated on the server to compute the optimal bin size using Sturges’ rule, resulting in 12 bins for the datasets used. We subsequently experimented with our binning strategies using this optimal value. *Fig 2* compares the hazard ratios (HR) of the clinical predictors and their 95% confidence intervals (CI) for both binning strategies relative to the standard federated Cox model.

**Fig 2.**
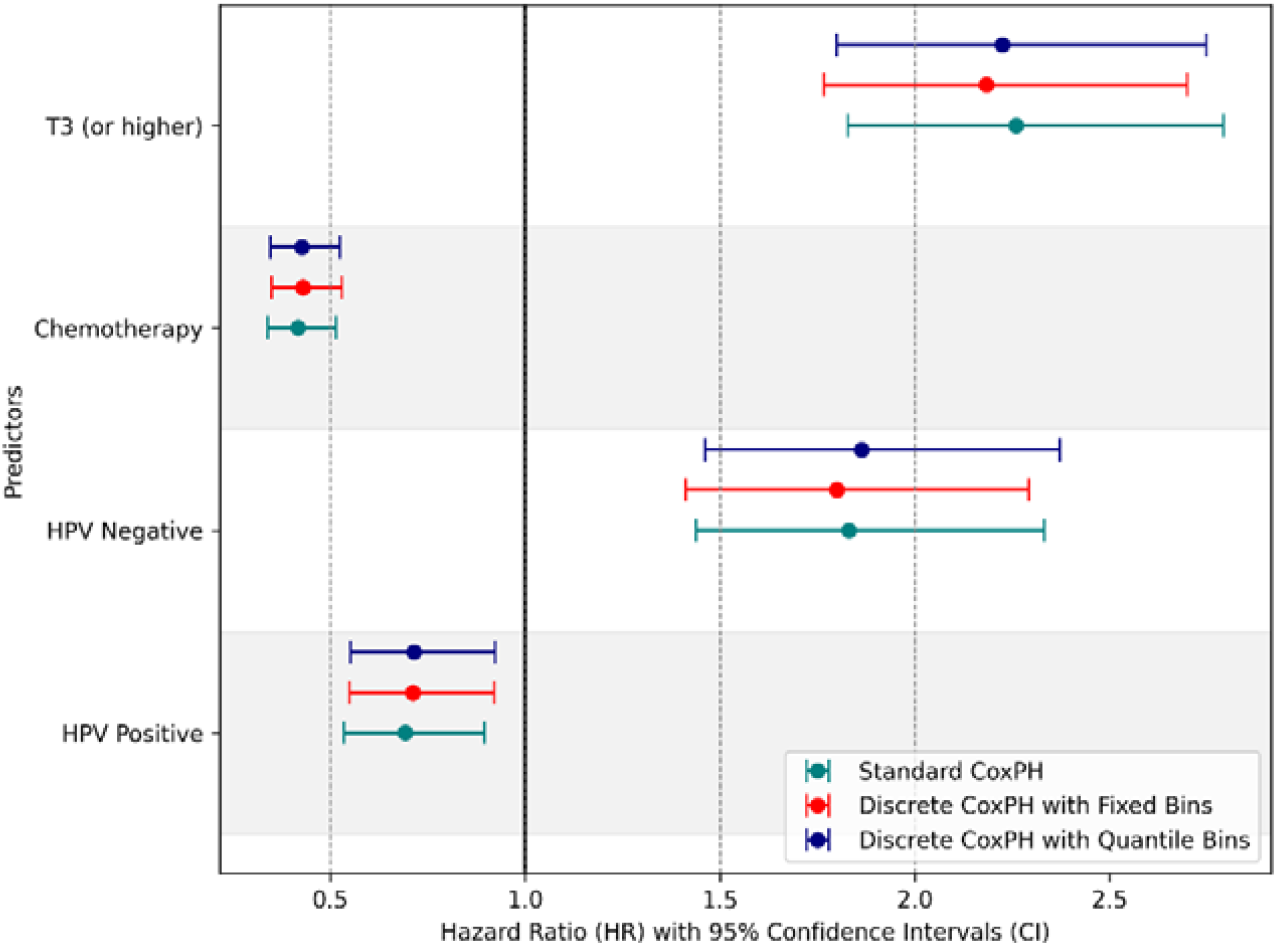
Comparison of Hazard Ratios (HR) with 95% Confidence Intervals (CI) of the clinical predictors for the two binning strategies relative to the standard federated Cox model.

**Fig 3.**
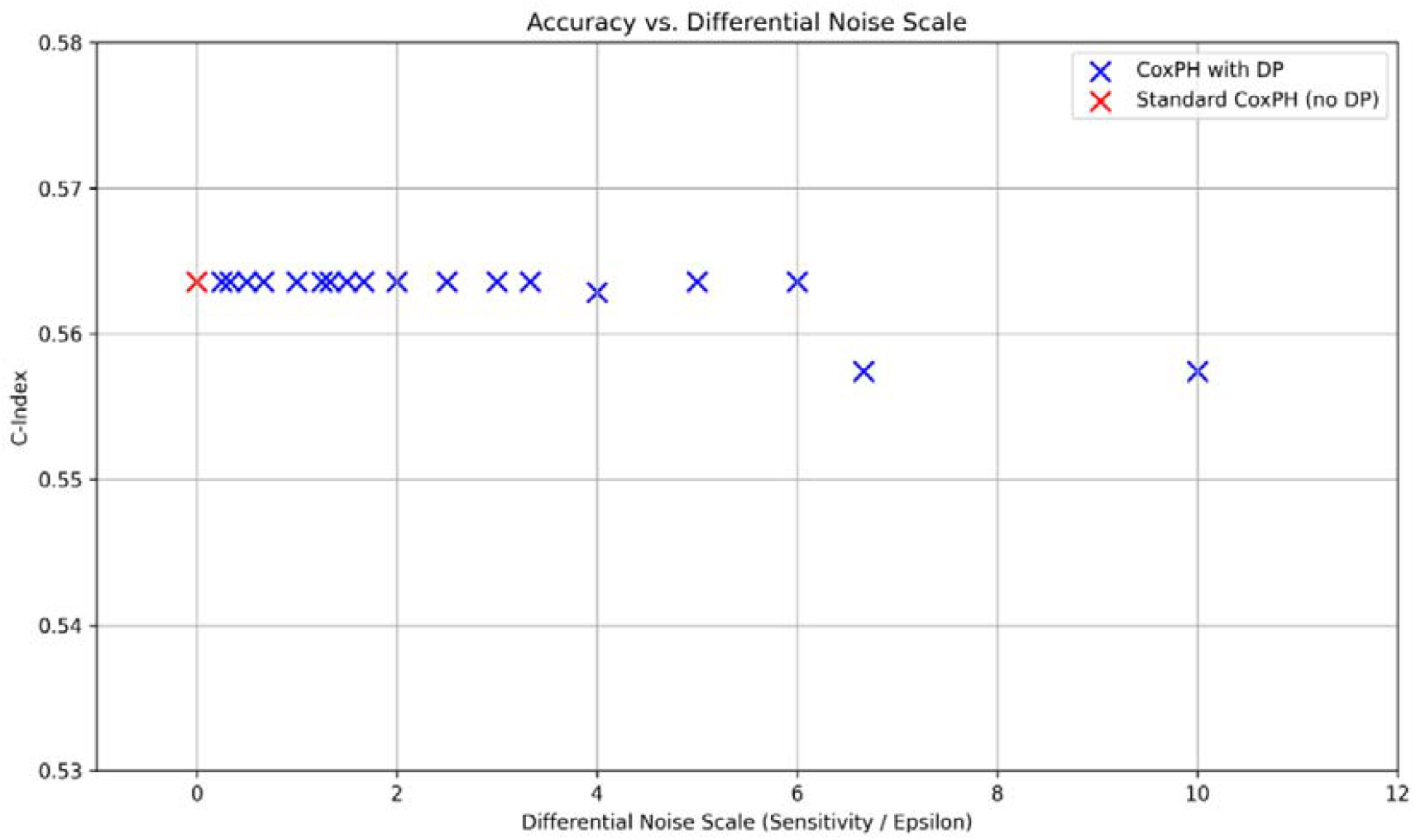
Accuracy (C-Indices) for different noise scales

### Differential Privacy on Partials

The results of our experiments to determine an optimal ε are presented in Fig 3. The C-Indices (shown in blue) remain relatively stable and comparable to the C-Index of the standard Cox model (shown in red) when the noise scale is low. However, beyond a certain threshold (around 4), the C-Index decreases, indicating a decline in model accuracy due to excessive noise. For our current models, we arbitrarily chose to investigate a noise scale of 3.3 (sensitivity = 1, epsilon = 0.3) to balance added noise and model accuracy.

We initially compared the outcomes of the model where noise was introduced to the partial results, specifically the partial gradients, before transmitting them to the server against those of the standard Cox model. In a follow-up analysis, noise was introduced to a 25% subset of input variables (predictors) at the local stations before computing the partial gradients. The HRs are presented alongside the standard Cox model results in *Fig 4* and *Fig 5*. Comprehensive results of the models can be found in the Supplementary Materials.

**Fig 4.**
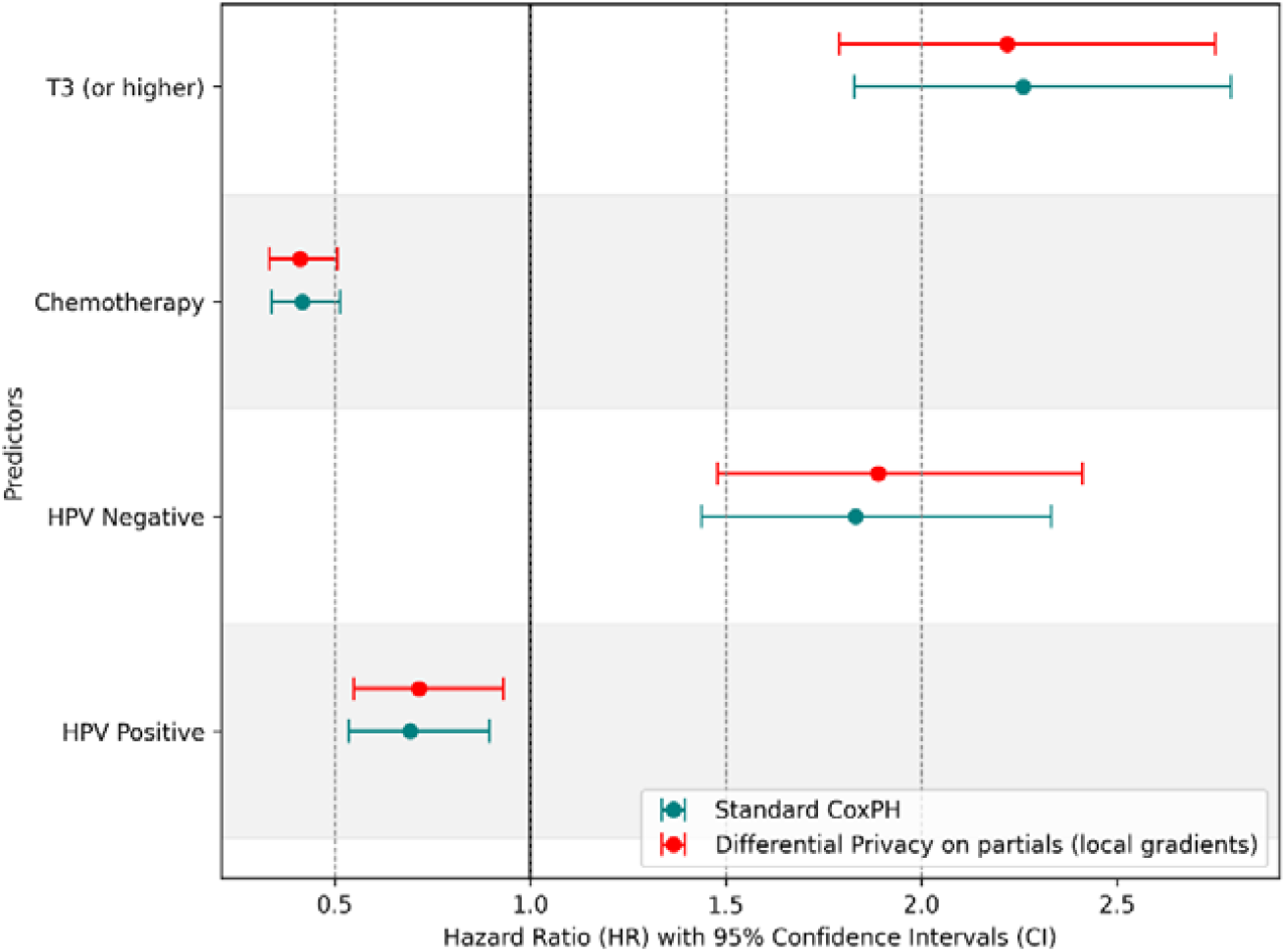
Comparison of Hazard Ratios (HR) with 95% Confidence Intervals (CI) of the clinical predictors when differential noise was added to partials relative to the standard federated Cox model.

**Fig 5.**
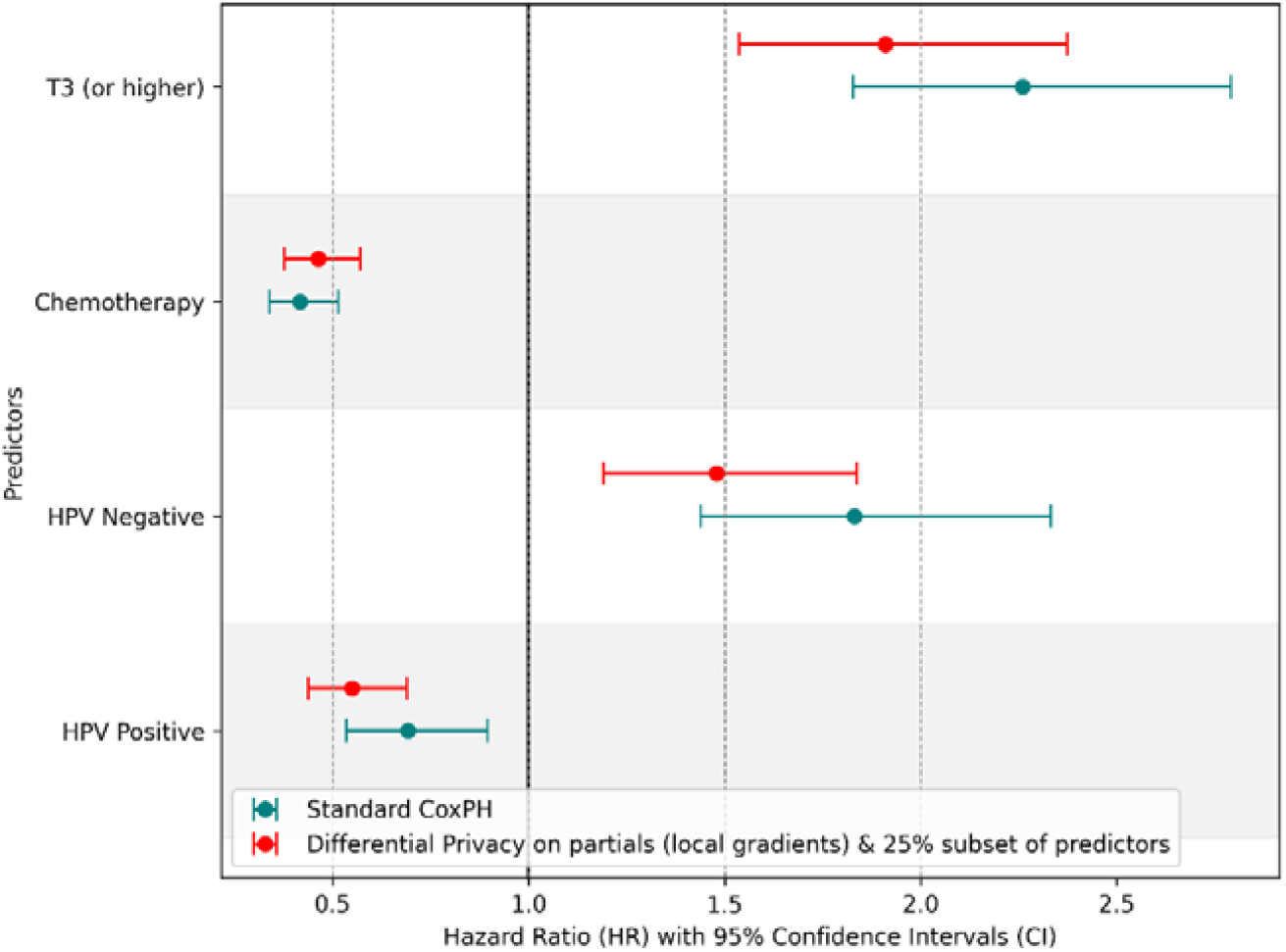
Comparison of Hazard Ratios (HR) with 95% Confidence Intervals (CI) of the clinical predictors when differential noise was added to partials and 25% subset of predictors relative to the standard federated Cox model.

### Cumulative Baseline Hazards

The cumulative baseline hazard curves for each strategy were individually derived from the Cox model results using the Breslow estimator. *Fig 6* shows the total number of patients at risk at each event time. The baseline hazard curves are presented in *Fig 7, 8, and 9*, representing event time binning, noise applied to the local gradients, and noise applied to both the predictors (25% subset) and gradients, respectively, for comparative analysis. *Fig 10* illustrates the differences in hazard values calculated in each PETs compared to the standard model.

**Fig 6.**
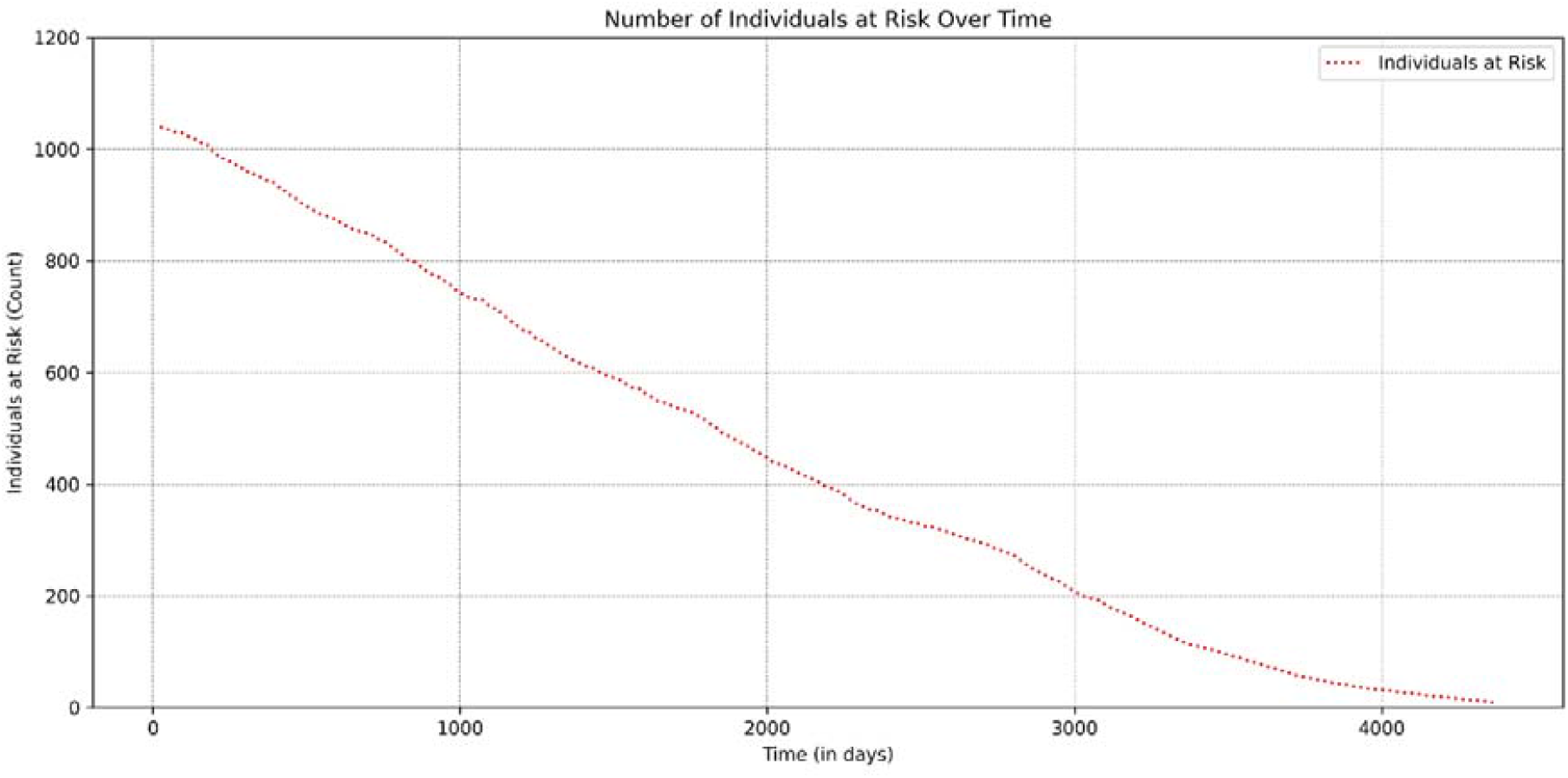
Total number of individuals at risk over time for all three datasets

**Fig 7.**
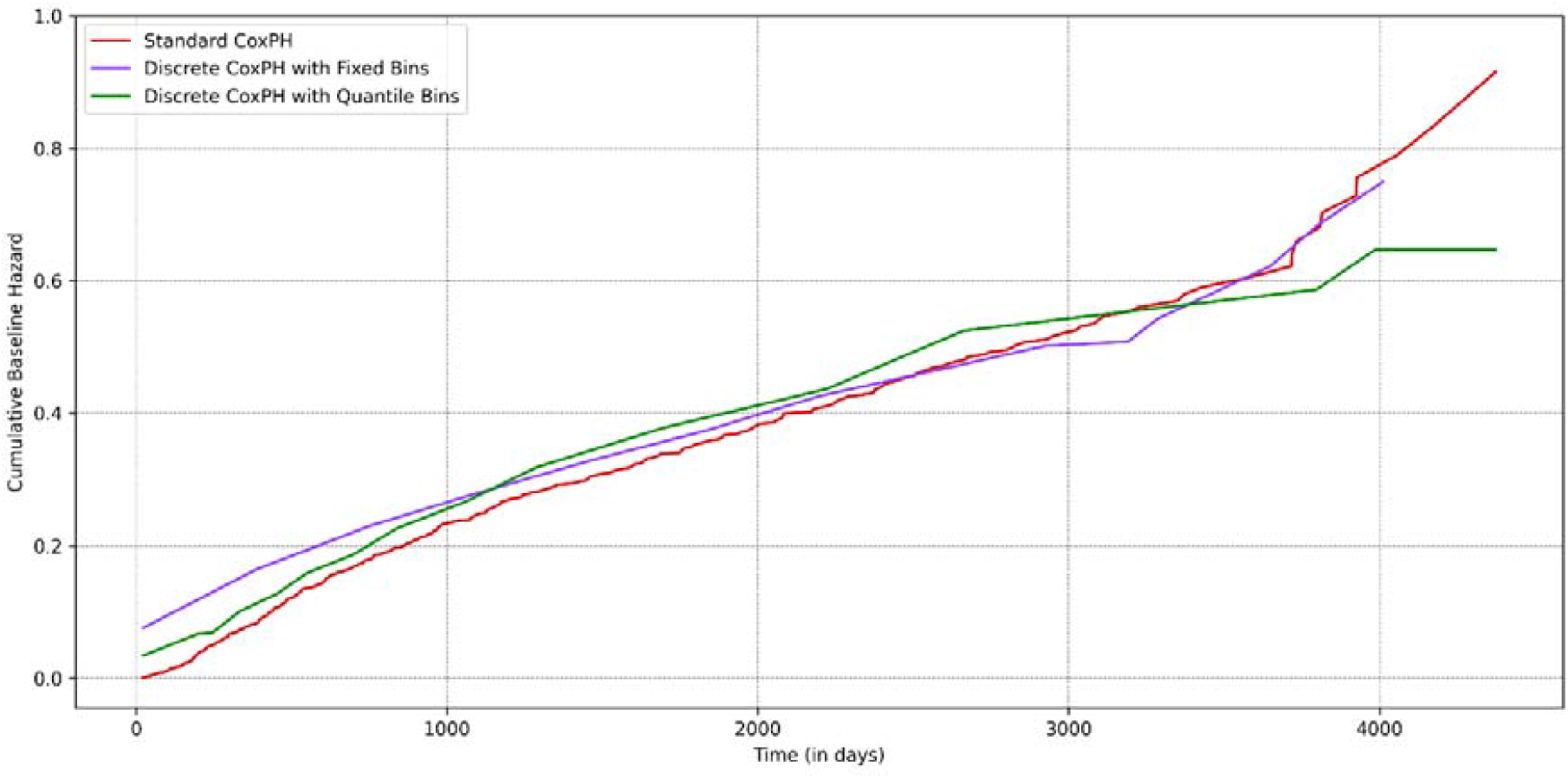
Cumulative baseline hazards for the two binning strategies relative to the standard Cox model.

**Fig 8.**
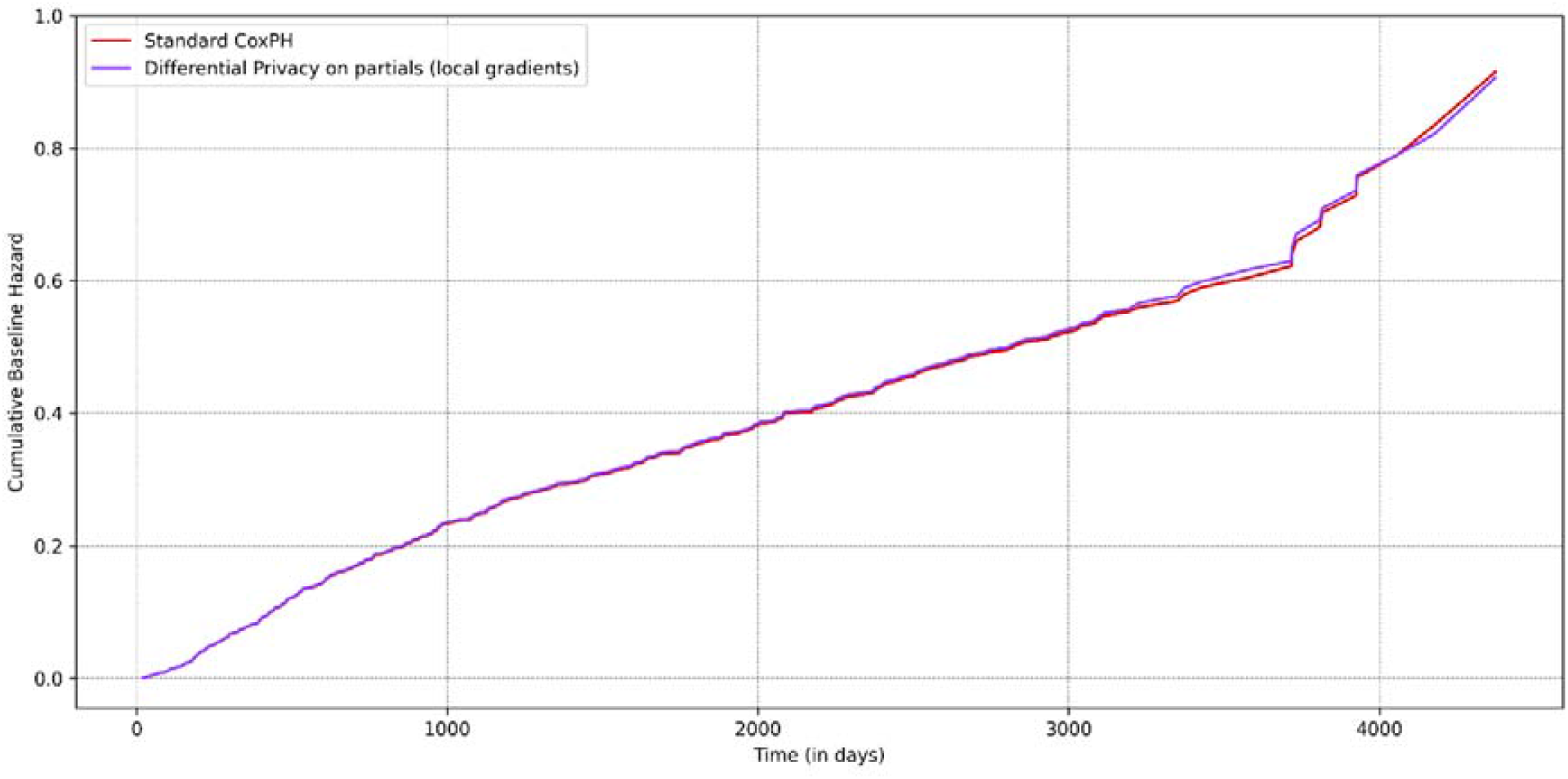
Cumulative baseline hazards when differential noise was added to partials, relative to the standard federated Cox model.

**Fig 9.**
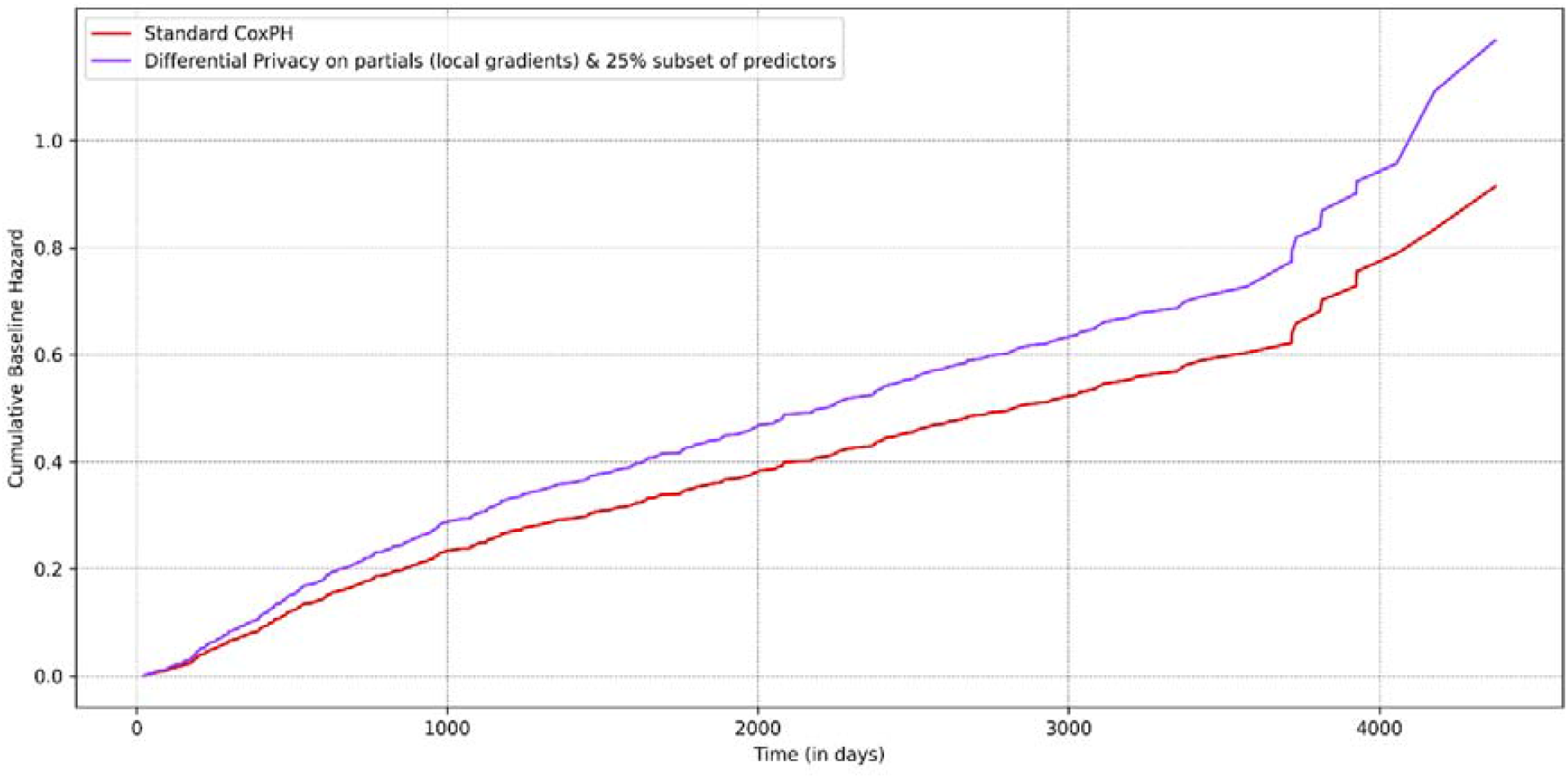
Cumulative baseline hazards when differential noise was added to partials and 25% subset of predictors, relative to the standard federated Cox model.

**Fig 10.**
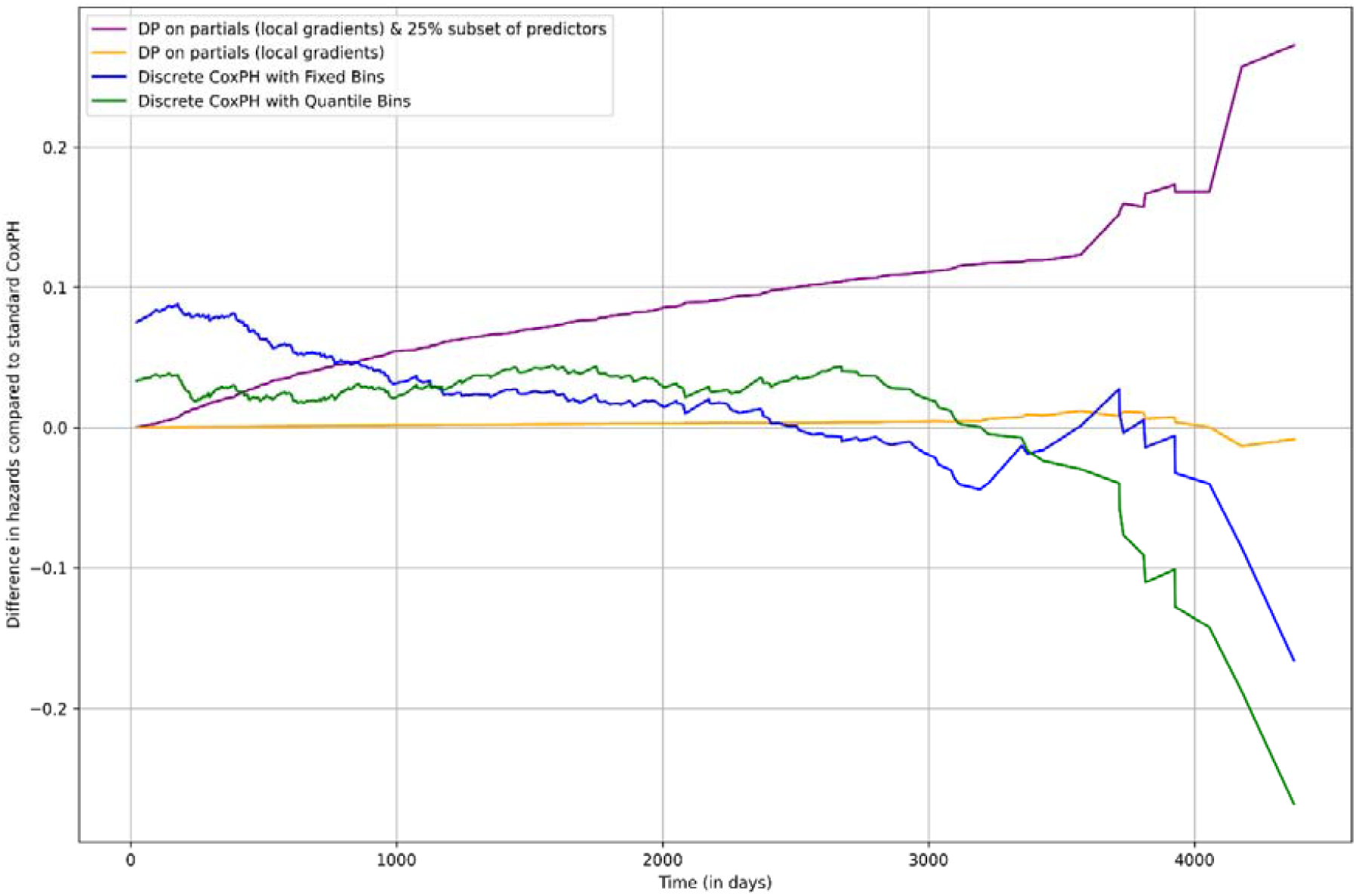
Differences in cumulative baseline hazards calculated for each privacy-enhancing technology (PET) relative to the standard federated Cox model.

## Discussion

In this study, we explored two potential enhancements to address possible data leakage issues in federated survival analysis when subjected to certain attacks, as was reported by recent research in the field. Our findings demonstrate that enhancing privacy in a federated learning environment is not only feasible but can also be tailored to meet project-specific requirements.

The Vantage6 software used in this work instantiates a PHT-type infrastructure with a client-server network topology, allowing for the analysis of multi-institutional datasets without the need to exchange sensitive patient-level data among collaborating parties. Instead, only aggregated cohort summaries or statistical model coefficients are shared through a mutually trusted third-party server, thus eliminating the need for direct peer-to-peer interactions between institutions. Moreover, Vantage6 supports web-industry-grade End-to-End (E2E) encryption. Even if an attacker were to intercept the transmissions between a local data station and the aggregator, it would remain indecipherable without the appropriate decryption keys, which are securely protected by the system.

We assessed two discretization strategies—Fixed and Quantile Bins—for additional privacy and compared them to traditional standard federated Cox models. Despite the binning implementation, the predictors’ HRs remained consistent across all models. This consistency indicates that the binned models maintain their predictive power with minimal effect on the overall associations within the data in the range of binning strategies we tested.

For the cumulative baseline hazard curves, fixed binning shows a steep initial rise, suggesting that while privacy is improved, there is a slight compromise in the hazard estimation. Meanwhile, the baseline curves for quantile binning closely align with those of the standard Cox model during the initial follow-up periods. However, some divergence is observed later, which may indicate increased sensitivity to data sparsity over time, as shown in *Fig 6*.

Experiments incorporating Differential Privacy (DP) into the Cox model yielded the following insights. Adding noise to the local gradients in the Cox model preserves the model’s utility while enhancing privacy. The HRs for the predictors remain consistent with those of the standard Cox model in our experiments, indicating that the noise added with the chosen epsilon value to the local gradients does not significantly alter the estimated effect sizes. Furthermore, the cumulative baseline hazard curves show minimal deviation between the standard model and the model with noise on local gradients, suggesting that the overall risk estimation remains reliable under this approach. When additional noise is applied to a 25% subset of the predictors along with the noise on local gradients, the results reveal a more pronounced divergence. This suggests that while the combination further enhances privacy, it may introduce a degree of overestimation in the baseline hazard values, especially over longer follow-up periods.

The epsilon parameter controls the trade-off between privacy and accuracy when incorporating DP. A smaller epsilon provides stronger privacy by introducing more noise but at the cost of compromising model performance. Conversely, a larger epsilon offers less privacy protection but retains more of the original data utility. In practice, choosing epsilon a priori depends on the sensitivity of the data. While the literature does not provide an ideal threshold for selecting epsilon to balance data utility and privacy, we propose the following for a systematic approach:

- **Initial testing**: Beginning with an intermediate epsilon value (e.g., 1.0 or 0.5) based on the project’s privacy requirements.
- **Sensitivity analysis**: Re-running the model with varying values of epsilon. For instance, in the case of survival analysis, if the HRs remain consistent across a range of epsilon (from 0.5 to 1.0), it indicates that model performance is largely unaffected by noise, and any value within this range is acceptable.
- **Divergence thresholds**: If significant deviations are observed in model outputs at lower epsilon values (e.g., ε < 0.5), it suggests that the noise is too disruptive. In such cases, reducing the noise may be necessary while still considering the acceptable level of privacy risk.

A Wald test was conducted to assess whether the models incorporating PETs yielded significantly different estimates for the effects of each predictor compared to the standard Cox model (*refer Table 1 for results*). The Wald statistics for the predictors in each PET model were relatively low, suggesting a likely non-significant difference. Moreover, the p-values were considerably above the 0.05 significance level, suggesting no statistically significant difference between the coefficients of the PET and standard models. These findings indicate that incorporating PETs in our Cox analysis resulted in comparable outcomes while preserving the overall predictive accuracy of the models. Although the experiments were not directly compared to a centralized Cox model, the standard federated Cox model used in this analysis was reported to produce similar results to that of a centralized analysis in the relevant literature [18].

**Table 1.** Wald Test Statistic results for assessing the statistical differences for each privacy-enhancing technology (PET) results relative to the standard Cox model.

| PET | Predictor | Wald Statistic | p-value |
| --- | --- | --- | --- |
| DP on Aggregates | Chemotherapy | 0.01 | 0.92 |
|  | HPV Negative | 0.03 | 0.86 |
|  | HPV Positive | 0.03 | 0.87 |
|  | T3 (or higher) | 0.01 | 0.91 |
| DP on Aggregates & 25% Predictors | Chemotherapy | 0.50 | 0.48 |
|  | HPV Negative | 1.67 | 0.20 |
|  | HPV Positive | 1.75 | 0.19 |
|  | T3 (or higher) | 1.18 | 0.28 |
| Fixed Bins | Chemotherapy | 0.04 | 0.84 |
|  | HPV Negative | 0.01 | 0.92 |
|  | HPV Positive | 0.02 | 0.88 |
|  | T3 (or higher) | 0.05 | 0.82 |
| Quantile Bins | Chemotherapy | 0.02 | 0.89 |
|  | HPV Negative | 0.01 | 0.92 |
|  | HPV Positive | 0.03 | 0.87 |
|  | T3 (or higher) | 0.01 | 0.92 |

In this work, we exclusively sampled noise from a Laplacian distribution for differential privacy. Alternatively, depending on the project’s preferences, one could opt to sample noise from a truncated Gaussian distribution. We experimented with adding Laplacian noise to 25% of the predictors and the partial results, increasing the complexity of data reconstruction in the event of a security breach. Applying noise to 100% of the predictors before computing local gradients would mean effectively transforming the data, making the computations resemble those performed on a completely different dataset. This can significantly alter the underlying relationships within the data, potentially resulting in biased estimates and misleading conclusions.

An alternative is synthetic data, which involves generating a new dataset that mimics the statistical properties of the original data. While both these methods aim to enhance privacy, they differ fundamentally in their implementation and implications. Synthetic data preserves the original data structure and relationships, whereas indiscriminately applying differential noise can obscure these relationships and degrade model performance. Researchers may opt for differential noise when strict privacy guarantees are essential [31], but they should be mindful of the potential loss of data utility. Synthetic data may be preferable when the objective is to maintain analytical utility while still protecting sensitive information. However, a synthetic data engine is a machine learning algorithm. Training such an algorithm requires access to the original data, which introduces its own privacy risks and – if learned too “ deep” – is not guaranteed to be privacy-preserving [32]. Also, the final version must be based on real-world data for trust in prediction models.

Another potential PET is using secure aggregation techniques such as Secure Multi-Party Computation (SMPC) [33], which ensures strong privacy protections by aggregating model updates without disclosing individual contributions. However, SMPC can be computationally demanding. In our study, we experimented with event time binning and differential privacy, which offer a balance between privacy and computational efficiency. These methods provide adequate protection against inference attacks while remaining practical for applications with limited resources and time.

Introducing binning strategies, particularly quantile-based approaches, offers a viable method for enhancing privacy in federated survival analysis. Future research could explore optimizing these binning strategies to minimize any observed trade-offs, such as the variability in cumulative hazard estimation, and to assess their performance across larger datasets. Improvements could also include incorporating outlier detection when calculating bin edges and setting a minimum threshold for the number of data points in each bin. Applying DP highlights the importance of carefully balancing the noise level with the need to maintain model accuracy (privacy vs utility), particularly in sensitive applications like federated survival analysis.

Selecting the appropriate PET is crucial and should be guided by the specific needs of the users and the data. In most federated modeling, End-to-end encryption (E2E) may provide sufficient privacy protection. However, in more sensitive cases where additional layers of privacy are necessary, both binning and DP offer valid options. Binning is more suited for defending against membership inference attacks, where the adversary tries to determine whether an individual is part of the dataset. On the other hand, DP helps protect against model inversion attacks, where the aim is to reverse-engineer individual data points using model gradients. For instance, in Cox analysis, researchers could begin with binning event times and then layer DP as required, adjusting the noise based on privacy requirements.

While the study demonstrates the feasibility of incorporating PETs into the federated Cox model, we also acknowledge the following limitations. Firstly, the experiments were conducted using public Head & Neck (HNC) datasets, which may not fully represent other datasets, particularly those with higher sensitivity. The equivalency of the privacy protections offered by our approach could be data-dependent. Different data distributions or patterns may introduce variations affecting the balance between privacy and utility, potentially leading to unreliable hazard estimates. Therefore, the generalizability of the PETs may vary when applied to different types of data, such as lung or prostate cancer datasets.

Secondly, we used the C-index to measure model performance while choosing the optimal epsilon value for the experiments. However, it is crucial to recognize that while the C-index may remain stable, the HRs could still vary, leading to inaccuracies in interpreting the effects of the predictors. Future studies might consider additional performance metrics or sensitivity analyses to ensure that noise introduced by DP does not mask critical shifts in HRs, which could influence clinical interpretations. Future work might also explore using PETs to protect more advanced analysis techniques, such as plotting Schoenfeld residuals to validate Cox model assumptions or constructing Kaplan-Meier curves within a federated framework.

## Conclusion

This study evaluated two PETs—event times binning and differential privacy (DP)—to mitigate data leakage risks in federated survival analysis. Using the Vantage6 infrastructure, we enabled secure multi-institutional analysis without sharing sensitive patient data. Both fixed and quantile binning strategies maintained consistent hazard ratios, although fixed binning showed slight distortions in cumulative hazard curves, indicating a minor trade-off between privacy and model accuracy. DP enhanced data security by adding noise to local gradients, preserving model fidelity, and minimizing privacy risks. However, adding noise to both gradients and a subset of predictors introduced some risk overestimation, highlighting the need to carefully calibrate privacy parameters to balance accuracy and privacy. Our findings suggest that these techniques can effectively enhance privacy in federated learning settings, but the choice of method should be tailored to specific project requirements. Future research aims to explore optimizing these strategies for larger datasets and in different applications.

## Data Availability

The public datasets used in this work - HN1 [23], HEAD-NECK [24], and OPC [25] are obtained from The Cancer Imaging Archive (TCIA). The dataset HN3 is not publicly available due to the data containing information that is considered privacy sensitive.

## Statements & Declarations

### Author Contributions

Varsha Gouthamchand (VG) was responsible for the python coding, statistical experiments, and setting up the Vantage6 infrastructure. Varsha Gouthamchand (VG) prepared most of the manuscript text under the supervision of Johan van Soest (JvS), Andrea Damiani (ADa), and Leonard Wee (LW). Johan van Soest (JvS), Giovanni Arcuri (GA), Andre Dekker (ADe), Andrea Damiani (ADa) and Leonard Wee (LW) jointly oversaw the overall supervision of the project. All authors reviewed the manuscript prior to submission and consented to its publication.

## Acknowledgments

None.

