## Supplementary Tables for "Navigating the Privacy-Accuracy Tradeoff: Federated Survival Analysis with Binning and Differential Privacy"

| Variable | Coef | Exp(coef) | SE | lower_CI | upper_CI | Z | p-value |
| --- | --- | --- | --- | --- | --- | --- | --- |
| T3 (or Higher) | 0.78102 | 2.18369 | 0.1081 | 1.76675 | 2.69904 | 10.95038 | <0.001 |
| Chemotherapy | -0.84406 | 0.42996 | 0.10641 | 0.34902 | 0.52967 | -5.35683 | <0.001 |
| HPV Negative | 0.58731 | 1.79914 | 0.12353 | 1.41226 | 2.29201 | 6.469226 | <0.001 |
| HPV Positive | -0.34058 | 0.71136 | 0.13129 | 0.54996 | 0.92012 | -2.19849 | 0.027 |

Table S1. Discrete CoxPH with Fixed bins

| Variable | Coef | Exp(coef) | SE | lower_CI | upper_CI | Z | p-value |
| --- | --- | --- | --- | --- | --- | --- | --- |
| T3 (or Higher) | 0.79927 | 2.22391 | 0.10796 | 1.79979 | 2.748 | 11.33695 | <0.001 |
| Chemotherapy | -0.85297 | 0.42615 | 0.10627 | 0.34602 | 0.52483 | -5.39974 | <0.001 |
| HPV Negative | 0.62232 | 1.86325 | 0.12345 | 1.46281 | 2.3733 | 6.992922 | <0.001 |
| HPV Positive | -0.33608 | 0.71457 | 0.13118 | 0.55256 | 0.92407 | -2.17593 | 0.029 |

Table S2. Discrete CoxPH with Quantile bins

| Variable | Coef | Exp(coef) | SE | lower_CI | upper_CI | Z | p-value |
| --- | --- | --- | --- | --- | --- | --- | --- |
| T3 (or Higher) | 0.79711 | 2.21912 | 0.10966 | 1.78993 | 2.75122 | 11.11728 | <0.001 |
| Chemotherapy | -0.8892 | 0.41099 | 0.10767 | 0.33279 | 0.50755 | -5.47065 | <0.001 |
| HPV Negative | 0.63596 | 1.88883 | 0.12455 | 1.4797 | 2.41109 | 7.136222 | <0.001 |
| HPV Positive | -0.33566 | 0.71487 | 0.13483 | 0.54885 | 0.9311 | -2.11473 | 0.034 |

Table S3. CoxPH with differential noise added on the local gradients

| Variable | Coef | Exp(coef) | SE | lower_CI | upper_CI | Z | p-value |
| --- | --- | --- | --- | --- | --- | --- | --- |
| T3 (or Higher) | 0.6473 | 1.91038 | 0.11087 | 1.53725 | 2.37407 | 8.21117 | <0.001 |
| Chemotherapy | -0.768 | 0.46394 | 0.10597 | 0.37693 | 0.57104 | -5.05878 | <0.001 |
| HPV Negative | 0.39171 | 1.47952 | 0.11023 | 1.19203 | 1.83631 | 4.35012 | <0.001 |
| HPV Positive | -0.59917 | 0.54927 | 0.11591 | 0.43764 | 0.68936 | -3.8885 | 0.0001 |

Table S4. CoxPH with differential noise added on the local gradients and 25% subset of predictors
